# Mask Interventions in K12 Schools Can Also Reduce Community Transmission in Fall 2021

**DOI:** 10.1101/2021.09.11.21263433

**Authors:** Jessica Mele, Erik Rosenstrom, Julie Ivy, Maria Mayorga, Mehul D. Patel, Julie Swann

## Abstract

The dominance of the COVID-19 Delta variant has renewed questions about the impact of K12 school policies, including the role of masks, on disease burden.^1^ A recent study showed masks and testing could reduce infections in students, but failed to address the impact on the community,^2^ while another showed masking is critical to slow disease spread in communities, but did not consider school openings under Delta.^3^ We project the impact of school-masking on the community, which can inform policy decisions, and support healthcare system planning. Our findings indicate that the implementation of masking policies in school settings can reduce additional infections post-school opening by 23-36% for fully-open schools, with an additional 11-13% reduction for hybrid schooling, depending on mask quality and fit. Masking policies and hybrid schooling can also reduce peak hospitalization need by 71% and result in the fewest additional deaths post-school opening. We show that given the current vaccination rates within the community, the best option for children and the general population is to employ consistent high-quality masking, and use social distancing where possible.

## INTRODUCTION

The dominance of the COVID-19 Delta variant has renewed questions about the impact of K12 school policies, including the role of masks, on disease burden.^1^ A recent study showed masks and testing could reduce infections in students, but failed to address the impact on the community,^2^ while another showed masking is critical to slow disease spread in communities, but did not consider school openings under Delta.^3^ We project the impact of school-masking on the community, which can inform policy decisions, and support healthcare system planning.

## METHODS

We use a stochastic agent-based Susceptible-Exposed-Infected-Recovered simulation model across a network representing North Carolina^4^ to simulate school-masking for K12 students and project infections, hospitalizations, and deaths. Each scenario begins July 1, 2021, with schools opening August 23, where the Delta variant is 93% dominant. We study scenarios with school-masking (100% compliance) or without (0% compliance) with schools fully open or hybrid (rotating half joining remotely). For students wearing masks, we vary mask efficacy (50% or 70% reduction in transmission and susceptibility^5^) to capture quality and fit. Beginning when schools open, we assume working adults wear masks (50% effective) at rates of 50%, 40%, or 30% for urban, suburban, and rural census-tracts, respectively. We incorporate age-based, county vaccination levels by July 1 and assume vaccinations continue at June 2021 rates with a maximum vaccination rate of 95%. The simulation results are validated on hospitalizations and deaths (July 1-August 23, 2021). We incorporate time-based reinfections and initialization of recovered/vaccinated by county and age (see supplement).

## RESULTS

Figure 1(a)-(d) shows cumulative infections by age. We find that when schools are open with no masks (blue-dashed line) the highest number of new infections occur in all age groups, with 80% more additional infections (occurring post-school opening) than the best scenarios studied.

**Figure 1 (a-d):**
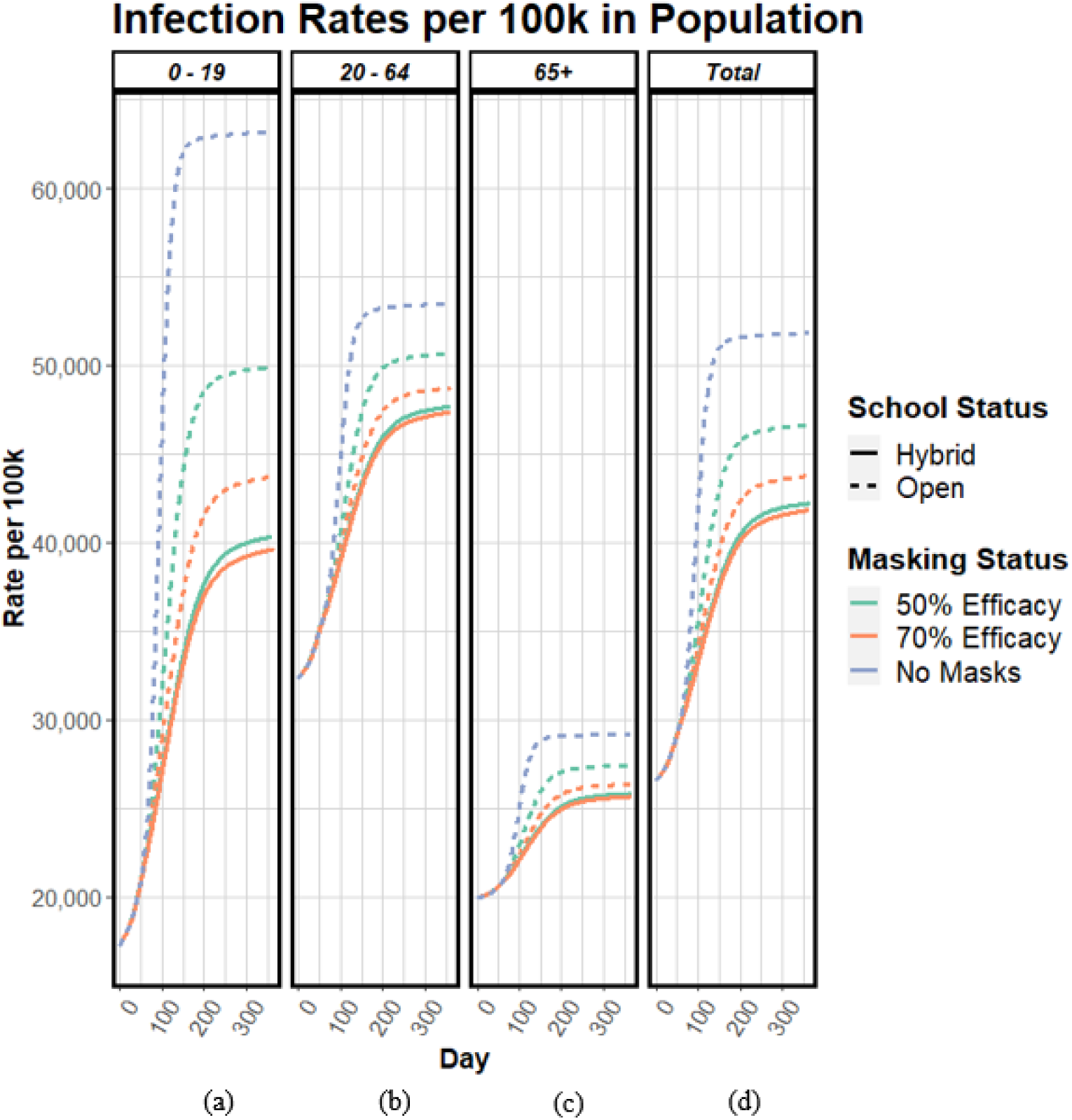
Cumulative Infections per 100k in Population by Age Group and Total. Note: The “No Masks” Scenario is only shown for open schools.

When students wear masks that are 50% effective we observe a 23% reduction in additional infections in the general population post-school opening, whereas we observe a 36% reduction in additional infections when students wear masks that are 70% effective, compared to no school-masking. Hybrid schooling provides an additional 11-13% reduction in additional cases, compared to the best school fully-open scenario.

Figure 2(a)-(b) displays hospitalizations and deaths over time. When schools are open without masking, the peak estimated hospitalization level exceeds the peak observed in January 2021, with over 18,000 new deaths occurring within 6 months of schools opening. If schools are hybrid or there is high mask-efficacy, we observe fewer deaths and a reduction in peak hospitalization need of 71%. With schools fully open, a 20% increase in school mask-efficacy leads to a 7% and 29% reduction in cumulative deaths and peak hospitalizations. With no school-masking, approximately 6%, 46%, and 48% of hospitalizations occur in children, adults, and 65+, respectively.

**Figure 2 (a,b):**
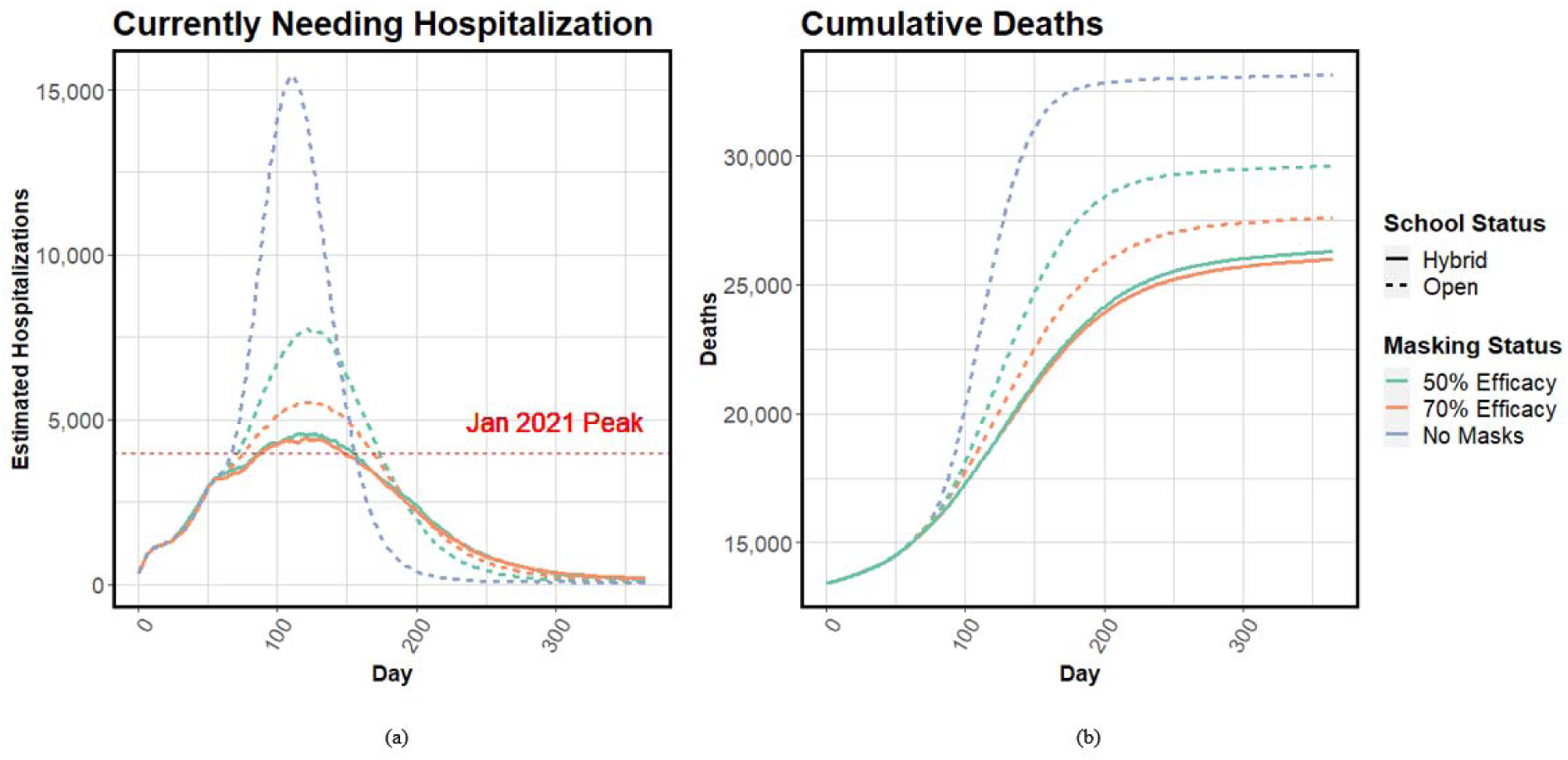
(a)Currently Needing Hospitalization and (b) Cumulative Deaths Over Time Notes: (a) The red-dashed, horizontal line signifies the highest observed number of hospitalizations as of September 10th, 2021, 3,992, in North Carolina since the start of the pandemic.

## DISCUSSION

We show school-masking can reduce infections in the overall community. Without school-masking, we show hospitals are likely to be further overwhelmed, which can increase all-cause mortality. Higher mask efficacy in schools can also reduce infections, hospitalizations, and deaths in the community. This supports ensuring all students consistently wear high-quality, well-fitting masks^6^. Increasing social distance (hybrid schooling) can further reduce negative outcomes although it is more disruptive to learning. The study did not include surveillance testing for further reductions, nor did it quantify school absences from isolation or quarantine, which is likely highest in no-mask scenarios.

We show that given the current vaccination rates within the community, the best option for children and the general population is to employ consistent high-quality masking, and use social distancing where possible.

## Data Availability

This manuscript uses publicly available data from the US Census, published estimates of disease parameters, data obtained from the North Carolina Department of Health Data Dashboard, and estimates obtained from the Covid-19 Case Surveillance Restricted Access Detailed Data.

## Author Contributions

Drs. Ivy, Mayorga and Swann had full access to all of the data in the study and take responsibility for the integrity of the data and the accuracy of the data analysis.

*Concept and design:* Mele, Rosenstrom, Ivy, Mayorga, Patel, Swann.

*Acquisition, analysis, or interpretation of data:* Mele, Rosenstrom Ivy, Mayorga, Swann, Patel.

*Drafting of the manuscript:* Mele, Rosenstrom, Swann

*Critical revision of the manuscript for important intellectual content:* All authors.

*Statistical analysis:* Mele, Rosenstrom, Ivy, Mayorga, Swann

*Administrative, technical, or material support:* Swann

## Disclaimer

The content is solely the responsibility of the authors and does not necessarily represent the official views of the NIH, CSTE, CDC, or the universities employing the researchers.

## Additional Contributions

Pinar Keskinocak

