## Supplemental Material for "Mask Interventions in K12 Schools Can Also Reduce Community Transmission in Fall 2021"

1. **SEIR Model**
   a. **Initialization:**

We use an extended SEIR to model the spread of infection due to the Sars-CoV-2 virus among a population of approximately 1 million agents, created to be representative of the population of North Carolina. To create the population, we compile various datasets at the census-tract level for the state. Upon initialization, each census tract in the model is scaled to be approximately 1/10th that of the reported Census^1^ population data, with agents who are each assigned an (i) age group, 0-9; 10-19; 20-64; 65+, and (ii) race/ethnicity group, Non-Hispanic, White; Non-Hispanic, Black; Hispanic; Non-Hispanic, Other, based on population estimates of each for a given tract. Additionally, we assign some agents to have a high-risk medical condition, which is determined from the state-wide prevalence of diabetes by age and race/ethnicity^2^.

We seed the model with current values on county-level deaths^3^, hospitalizations^4^, and estimated recoveries and current active infections^3^. We estimate recovered and active infections using a time-varying lab multiplier, which is estimated from applying a population- average infection fatality ratio, obtained from age-based infection fatality ratios defined in the literature^5^ and the distribution of lab-confirmed cases in North Carolina^6^, prior to vaccine distribution, i.e. 2020 and for January-June 2021. As seen in

eTable1, we estimate lab multiplier values for 2021 through a monthly, linear decrease in infection fatality ratios by age from January 2021 through July 1st, 2021 dependent upon the proportion of the population that had been fully vaccinated. The seeding mechanism and lab multiplier is further defined in the Supplement to a previous study^7^.

|  | **2020** | **Jan-21** | **Feb-21** | **Mar-21** | **Apr-21** | **May-21** | **Jun-21** |
| --- | --- | --- | --- | --- | --- | --- | --- |
| **0-9** | 0.00001 | 0.00001 | 0.00001 | 0.00001 | 0.00001 | 0.00001 | 0.00001 |
| **10-19** | 0.00003 | 0.00003 | 0.00003 | 0.00003 | 0.00003 | 0.00003 | 0.00003 |
| **20-29** | 0.00011 | 0.00011 | 0.00011 | 0.00011 | 0.00011 | 0.00011 | 0.00011 |
| **30-39** | 0.00034 | 0.00034 | 0.00034 | 0.00034 | 0.00034 | 0.00034 | 0.00034 |
| **40-49** | 0.00114 | 0.00114 | 0.00114 | 0.00114 | 0.00114 | 0.00114 | 0.00114 |
| **50-59** | 0.00382 | 0.00382 | 0.00382 | 0.00382 | 0.00382 | 0.00382 | 0.00382 |
| **60-69** | 0.01277 | 0.01129 | 0.00867 | 0.00565 | 0.00303 | 0.00127 | 0.00039 |
| **70-79** | 0.04274 | 0.03778 | 0.02902 | 0.01892 | 0.01014 | 0.00426 | 0.00129 |
| **80+** | 0.14301 | 0.12642 | 0.09709 | 0.06330 | 0.03393 | 0.01425 | 0.00433 |
|  | Levin | Adjusted Levin | | | | | |

**eTable1:** Age-Based Infection Fatality Ratios with Adjustments post-vaccine distribution

b. **Disease Progression**

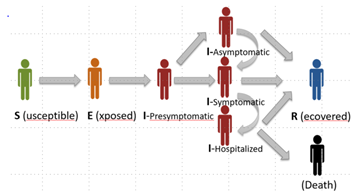

**eFigure1:** Overview of Disease States and Transition Directions
Source: Rosenstrom E, et al. High-Quality Masks Can Reduce Infections and Deaths in the US. *Forthcoming in Winter Simulation Conference Proceedings.* **doi:** https://doi.org/10.1101/2020.09.27.20199737^8^.

Agents in the model can exist in one of eight disease states (eFigure1), (i) Susceptible, (ii) Exposed, (iii) Presymptomatic Infected, (iv) Asymptomatic Infected, (v) Symptomatic Infected, (vi) Hospitalized, (vii) Dead, (viii) Recovered. The transition from one disease state to another is governed by probability distributions which are dependent upon an agent's age and status of high-risk medical conditions. An agent with diabetes is more at-risk for severe disease, modeled with a hospitalization rate that is three times larger than the rate for non-diabetic agents in the model^9^.

c. **Validation**

The model is validated through August 23, 2021 on hospitalizations reported by the North Carolina Department of Health and on deaths reported by the New York Times. The model has been similarly validated for other studies related to COVID-19 disease spread, interventions, and vaccine distribution^7,8,10,11^.

2. **Vaccinations**

Vaccinations in the model function by moving an agent directly to a Recovered disease state. We assume vaccines are 50% and 88% effective^12^, for one dose and two doses, respectively. We seed the model with current values of fully effective, fully vaccinated individuals at the county-level, by age groups.

We model continued vaccination distribution through the end of the simulation by county and age dependent upon the rate of vaccine uptake within each population from June to July, 2021. Since there is limited information about the rate of vaccinations in children, we assume that children will follow the same uptake patterns as the adults within their counties and allow the population to reach 75% of the proportion of total adults vaccinated within each county by the end of the simulation period. For counties with poor data reporting in which the populations of fully vaccinated 65+ reach above 100% with publicly available data^4^, we assume that the populations begin with 90% fully vaccinated upon seeding, and reach 95% fully vaccinated by the end of the simulation.

3. **Variants**

We model the Delta variant through a linear increase in the overall transmissibility of the virus over the first four weeks of the simulation. We modify the governing transmissibility to be consistent with the weighted average of observed strain types, e.g., Alpha, Delta, and wild types within the population. We assume that the Alpha variant is 20% more transmissible than the wild type and the Delta variant is 60% more transmissible than the Alpha variant^13^. That is, the estimated effective reproductive number increases from approximately 3.385 to 4.47 when Delta comprises 36.5% to 93% of the observed cases from early July to the end of July, respectively^14^.

4. **Reinfections**

To capture waning immunity, breakthrough infections, and reinfections, agents return to the Susceptible disease state at rates of 12% and 16%^15^, for those who entered the Recovered state from being fully vaccinated and infection, respectively. For agents who enter the Recovered state during the simulation, there is a base immunity of 6 months before they return to the Susceptible state. Those who enter the Recovered state during seeding either have a base immunity of 90 days if they are Recovered from infection, or 30 days for 65+, or 60 days for adults, if they are Recovered from being fully vaccinated. These numbers allow agents to return to the Susceptible state approximately 8 months after each age group was eligible for vaccination during the priority group phases in North Carolina^16^.

If an agent is selected to become reinfected, either from vaccination or previous infections, the respective probability of becoming infected with symptomatic disease is 0.36^17^, or ~50% lower than an agent with no history of immunity. We estimate increased hospitalization rates based on various datasets, and the calibration of the model to match observed data during the validation timeline^3,4,6,7^. There are no further adjustments made for probabilities of an agent to develop severe disease. The remaining disease parameters can be found in the Supplement of a previous study^7^.

| **Age Group** | **Hospitalization Rates** |
| --- | --- |
| 0-19 | 0.008 |
| 20-64 | 0.05 |
| 65+ | 0.3 |

**eTable2**: Hospitalization Rates by Age Group, Model Parameters^3,4,6,7^

3.	Coronavirus (Covid-19) Data in the United States. <https://github.com/nytimes/covid-19-data>
